# The ratio of BRAFV600E alleles can be used to assess the biological behavior of papillary thyroid carcinoma

**DOI:** 10.1101/2020.10.31.20223610

**Authors:** Dingcun Luo, Yeqin Ni, Shirong Zhang, Yanping Xun, Pan Zhao, Fan Wu, Tianhan Zhou, Jingjing Shi, Si Lu, Sihan Sun, Kaining Lu

## Abstract

**Background:** The BRAFV600E mutations is an important molecular event in the occurrence and development of papillary thyroid carcinoma (PTC). A qualitative detection of the BRAFV600E mutation is still insufficient to explain the biological behavior of PTC. Though quantitative detection of the BRAFV600E mutation can reflect certain characteristics of PTC, its clinical value is still controversial. We aimed to investigate the association between the ratio of BRAFV600E alleles and clinicopathological parameters in PTC patients.

**Methods:** Genomic DNA was extracted from specimens obtained from 329 PTC patients undergoing thyroidectomy. The ratio of BRAFV600E alleles was determined by amplification refractory mutation system (ARMS) and droplet digital polymerase chain reaction (ddPCR). Inconsistent results were further verified by next-generation sequencing (NGS). The clinicopathologic features, clinical tumor stage, and tumor recurrence risk stratification of all patients were correlated with the ratio of BRAFV600E alleles.

**Results:** The sensitivity of ddPCR was superior to that of ARMS and almost the same as that of NGS. In total, 275 of 329 patients had the BRAFV600E mutation as determined by ARMS, ddPCR and NGS. The ratio of BRAFV600E alleles ranged from 0.17%-48.0%, with a median ratio of 12.58%, and significantly correlated with tumor size (p<0.001), capsule or extrathyroidal invasion (p<0.001), the number or rate of lymph node metastases (p<0.001), tumor stage (p=0.006) and tumor recurrence risk (p<0.001) but not with sex, age or multifocality. The ratio of BRAFV600E alleles was much lower in PTC patients with Hashimoto’s thyroiditis than in those without (p<0.001).

**Conclusions:** The ratio of BRAFV600E alleles can reliably reflect the biological behavior of PTC, making it a molecular-based stratification index of recurrence risk. The quantitative detection of BRAFV600E has the potential to guide the clinical diagnosis and treatment of PTC.

## INTRODUCTION

Papillary thyroid carcinoma (PTC) is the most common malignant tumor in the endocrine system, and its incidence has rapidly increased in the past decade(1). Although most PTCs progress slowly, with a high survival rate and a low recurrence rate, some patients are very prone to cervical lymph node metastasis (LNM) and distant metastasis and even die of persistent or recurrent disease that is refractory to conventional therapies(2-4). Large-scale multicenter studies have demonstrated an association between BRAFV600E and PTC prognosis as well as PTC-specific mortality(5). Data from meta-analyses have consistently indicated that the presence of the BRAFV600E mutation in PTC is associated with extrathyroidal invasion, lymph node metastasis, advanced TNM stage, and tumor recurrence(6-9). Interestingly, PTCs with aggressive biological behaviors, such as those with the BRAFV600E mutation, can occur in small tumors, and thyroid microcarcinomas with the BRAFV600E mutation are frequently associated with extrathyroidal extension and lymph node metastasis(10). BRAF is a cytoplasmic serine-threonine protein kinase. Among the three forms of RAF kinases, BRAF is the most potent activator of the mitogen activated protein kinase (MAPK) pathway. The vast majority of BRAF allele alterations are characterized by a single amino acid substitution of valine by glutamic acid in a mutational hotspot at amino acid position 600 (BRAF c.1799T>A (p.Val600Glu), referred to hereafter as BRAFV600E(11-13). The BRAFV600E mutation is a negative prognostic marker in PTC.

With the development of detection techniques including amplification refractory mutation system (ARMS) and next-generation sequencing (NGS), the BRAFV600E mutation can be found in approximately 79%-87% of PTC patients, and it is an important molecular marker for PTC diagnosis, treatment and prognosis(12-18). The significance of the BRAFV600E mutation in the qualitative diagnosis of thyroid nodules is increasing, but the association between the BRAFV600E mutation and clinicopathological features and its impact on clinical outcomes remain challenging or unclear in PTC, especially in patients with microcarcinoma(19, 20). Guerra et al. demonstrated that completely clonal PTC harboring the BRAFV600E mutation was rare and that PTC was frequently associated with a mixture of wild-type and mutant BRAF alleles. The association between the BRAFV600E mutation and the clinicopathological features of PTC should be reevaluated, taking into account the fraction of mutant BRAF alleles(21, 22). We speculated that the larger the proportion of mutated BRAF alleles detected, the more aggressive the tumor. Therefore, the quantitative detection of BRAFV600E alleles should help clarify the relationship between the BRAFV600E mutation and the clinical behavior of PTC patients.

Droplet digital PCR (ddPCR) is a novel technology that provides sensitive and absolute nucleic acid quantification. It is far more sensitive and specific than other methods because of amplification at the nanoliter level and can accurately measure trace amounts of nucleic acids. Detection of the BRAFV600E mutation by ddPCR has been demonstrated in melanoma samples and pancreas fine needle aspiration (FNA) specimens(23-25). A recent study showed that mutant BRAFV600E was detected in 56%(42/75) of PTC tumors by ddPCR and that the ratio of mutant/total BRAF alleles varied from 4.7% to 47.5%, suggesting that ddPCR detection of the BRAFV600E mutation should be a suitable assay for the molecular-based stratification of prognosis in patients with PTC(26).

According to the 2015 American Thyroid Association (ATA) Management Guidelines for Differentiated Thyroid Cancer on the risk stratification of PTC recurrence(27), in this study, we investigated a correlation between the ratio of BRAFV600E alleles and disease risk (high, intermediate and low). We aimed to provide a reliable and molecular-based risk stratification for PTC patients. To this end, we used ddPCR and ARMS to detect the ratio of BRAFV600E alleles in PTC tissues and paracancerous tissues. If the results of ddPCR and ARMS were inconsistent, NGS was used for further clarification.

## MATERIALS AND METHODS

### Patients and sample correction

A total of 329 PTC patients who visited Affiliated Hangzhou First People’s Hospital, Zhejiang University School of Medicine between 2016 and 2018, and their matched formalin-fixed paraffin-embedded (FFPE) blocks were randomly and consecutively selected and analyzed for the BRAFV600E mutation. The exclusion criteria were as follows: (1) patients who had a history of treatments for thyroid cancer, (2) patients with other cervical malignant tumors, and (3) patients with both thyroid cancer and Graves’ disease or other endocrine disorder. Fresh tumor tissues and their paracancerous tissues were carefully dissected during the operation in 10 of 329 PTC patients. PTC tissues were collected from the center of the tumor lesions, and paracancerous tissues were obtained at least 2 cm from the margins of the tumor lesions. Histological diagnoses were established according to the WHO classification after the examination of hematoxylin and eosin-stained slides. The pathological diagnosis was performed by two thyroid pathologists. If there were different opinions, a third expert participated in the diagnosis, and the opinions of the two experts were used as the final diagnosis. A total of 144 PTC patients underwent total thyroidectomy, and 185 underwent hemithyroidectomy. All patients underwent neck central compartment lymph node dissection. Fifty-seven patients underwent therapeutic neck lateral compartment lymph node dissection. In this study, we extracted the following clinicopathologic variables: sex, age, multifocality, tumor diameter, capsule invasion, extrathyroid extension, Hashimoto’s thyroiditis (HT) and lymph node metastasis. Each patient was classified according to the 8th revision of the TNM staging system by the American Joint Committee on Cancer(AJCC)(28). The diagnostic criteria of Hashimoto’s thyroiditis were based on pathology or serum thyroid globulin antibody and peroxidase antibody combined with ultrasound characteristics(29,30). According to the risk of structural disease recurrence offered by the 2015 ATA Management Guidelines for Differentiated Thyroid Cancer, all patients were stratified as low, intermediate and high risk(27). All patients were followed up for a period of 16-50 months. One patient died of lung metastasis, and another patient developed recurrent cervical lymph node metastases. The remaining patients survived without recurrence. This study was approved by the Institutional Ethics Committee of Affiliated Hangzhou First People’s Hospital, Zhejiang University School of Medicine. All patients provided informed consent, and they all agreed to participate in this study. All procedures were performed in accordance with the Declaration of Helsinki.

### DNA extraction

The extraction of DNA from fresh frozen thyroid tissue was performed using a QIAamp DNA Mini Kit (QIAGEN Inc., Hilden), following the manufacturer’s instructions. The FFPE blocks of PTC samples were deparaffinized with xylene and rehydrated through ethanol (100%). Then, the extraction of DNA was performed using a QIAamp DNA FFPE Tissue Kit (QIAGEN Inc., Hilden) according to the manufacturer’s instructions. DNA quantification was performed on a NanoDrop 1000 (Thermo Fisher Inc., Waltham) according to the manufacturer’s instructions. The DNA had a purity 1.8 ≤A_260_/ A_280_ ≤ 2.0 and a concentration greater than 10.0 mg/L.

### ddPCR

ddPCR was performed on a QX-200 Droplet Digital PCR System (Bio-Rad Inc., Hercules). The primer and probe for detection of the BRAFV600E mutation were purchased from Bio-Rad. Two probes with one nucleotide difference that target the mutated region were labeled with carboxyfluorescein (FAM) and green fluorescent protein (VIC) dyes to detect mutant and wild-type BRAFV600E alleles, respectively. The final TaqMan PCR mixture (20 μL) containing 50 ng of template DNA was prepared. The thermal cycling conditions for the BRAFV600E detection assay comprised a 10-min incubation at 95 °C followed by 40 cycles at 94 °C for 15 s and 58 °C for 1 min, followed by a hold step at 4 °C for 5min. The analyses of ddPCR data for allele calling was performed with Quanta Soft software version 1.3.2.0 (Bio-Rad). All experiments and data analysis were performed according to the manufacturer’s instructions (YuanQi BioTech, China). Considering that single, nonspecific droplets were occasionally found in the positive area, the presence of at least three droplets with the FAM signal was defined as positive signal for the mutation. The ratio of BRAFV600E mutations was calculated with the following equation: (FAM positive droplet + FAM/VIC dual positive droplet)/(FAM positive droplet + VIC positive droplet + FAM/VIC dual positive droplet). The threshold was set at 0.1% of the ratio, as described in the manufacturer’s instructions.

### ARMS

ARMS was performed using an ABI 7500 system (Thermo Fisher Inc., Waltham). The ARMS kit for BRAFV600E mutation detection was purchased from AccBio Inc. (Beijing). The final PCR mixture (45 μL) was amplified under the following conditions: 1 cycle at 95 °C for 5 min; 15 cycles at 95 °C for 25 s and 64 °C for 20 s, followed by 72 °C for 20 s; and 31 cycles at 93 °C for 25 s, 60 °C for 35 s and 72 °C for 20 s. The FAM signals were detected at 60 °C. The definition of BRAFV600E status was classified by the FAM cycle threshold(Ct) value: (1) if the Ct value was <35, the sample was defined as positive; (2) if the Ct value was between 35-38, the sample was defined as suspicious; and (3) if the Ct value was >38, the sample was defined as negative.

### NGS

Due to the inconsistent results produced by ddPCR and ARMS, 16 of 329 specimens were analyzed with an AmpliSeq Cancer Hotspot Panel (v2) (Life Technologies Inc., Carlsbad) as described previously(31). BRAFV600E sequencing data were analyzed using Torrent Suite (Life Technologies Inc., Carlsbad). Mutations were identified and annotated through both Torrent Variant Caller and a direct visual inspection of the binary sequence alignment/map (BAM) file on the Broad Institute’s Integrative Genomics Viewer (IGV) (32).

### Normalization of BRAFV600E alleles to the proportion of neoplastic cells

The ratio of BRAFV600E alleles obtained by ddPCR was normalized to the proportion of neoplastic cells (vs contaminant cells, such as inflammatory or stromal cells) in each sample using the following formula: (normalized ratio of BRAFV600E alleles = ratio of BRAFV600E alleles / proportion of neoplastic cells). The proportion of neoplastic cells was calculated as a percentage after a careful microscopic evaluation of the tumor area on the hematoxylin and eosin-stained slide by two pathologists separately.

### Statistical Analysis

Medians with interquartile ranges were calculated for continuous variables, and numbers and percentages were calculated for categorical variables. Normality was tested with Kolmogorov-Smirnov tests. The Mann-Whitney *U* test was used to compare continuous, nonnormally distributed variables. The Kruskal-Wallis H test was used to compare variables between more than two groups. Pearson’s chi-square test or Fisher’s exact test was used to compare categorical variables. The Spearman rank-correlation test was used to analyze the correlation between two continuous variables. All statistical analyses were performed with SPSS statistical software (version 25.0). A *P* value<0.05 was considered statistically significant.

## Results

### Detection of the BRAFV600E mutation by ddPCR, ARMS and NGS

Among the 329 patients, 274 (83.28%) were detected the BRAFV600E mutation by ddPCR, and 263 (79.94%) were found to have the BRAFV600E mutation by ARMS. There was no significant difference between the data obtained from the two methods (p=0.268, Table 1). The ARMS and ddPCR results were inconsistent for 16 patients; therefore, NGS was conducted. The results obtained from NGS were consistent with those obtained from ddPCR for 15 patients and with those obtained from ARMS for one patient (Supplemental table 1). Consistent results between the two methods were considered the final result, revealing that 275 of 329 patients had the BRAFV600E mutation (83.58%).

**Table 1.** The positive rate of BRAFV600E mutation detected by ddPCR and ARMS in PTC.

| | BRAF-positive | BRAF-negative | $\chi^2$ | *P value* |
| --- | --- | --- | --- | --- |
| ddPCR | 274 ( 83.28% ) | 55 ( 16.72% ) | 1.225 | 0.268 |
| ARMS | 263 ( 79.94% ) | 66 ( 20.06% ) |  |  |

In 274 of 275 patients with the BRAFV600E mutation as determined by ddPCR, the ratio of BRAFV600E alleles varied from 0.17% to 48.0% (mean 15.79%), the quartiles were 6.3% and 22.71% (median 12.58%). If the ratios of BRAFV600E alleles obtained by ddPCR were divided into three groups, >10%, 1-10%, and <1%, the positive rates of BRAFV600E mutations detected by ARMS were 98.1%, 94.1%, and 69.2%, respectively (Figure 1). Although there was no statistically significant difference in the sensitivity of ddPCR versus that of ARMS, the sensitivity of BRAFV600E mutation detection by ddPCR was notably higher than that of ARMS. Additionally, analysis by ddPCR indicated that there was a difference in the average ratio of BRAFV600E alleles between 10 fresh frozen tissues and 10 FFPE-matched specimens (30.40% vs. 23.00%). However, paraffin tissues were utilized uniformly for analysis in the present study because of the statistically nonsignificant difference (P=0.302). No BRAFV600E mutation was found in 10 thyroid paracancerous tissue specimens.

**Figure 1.**
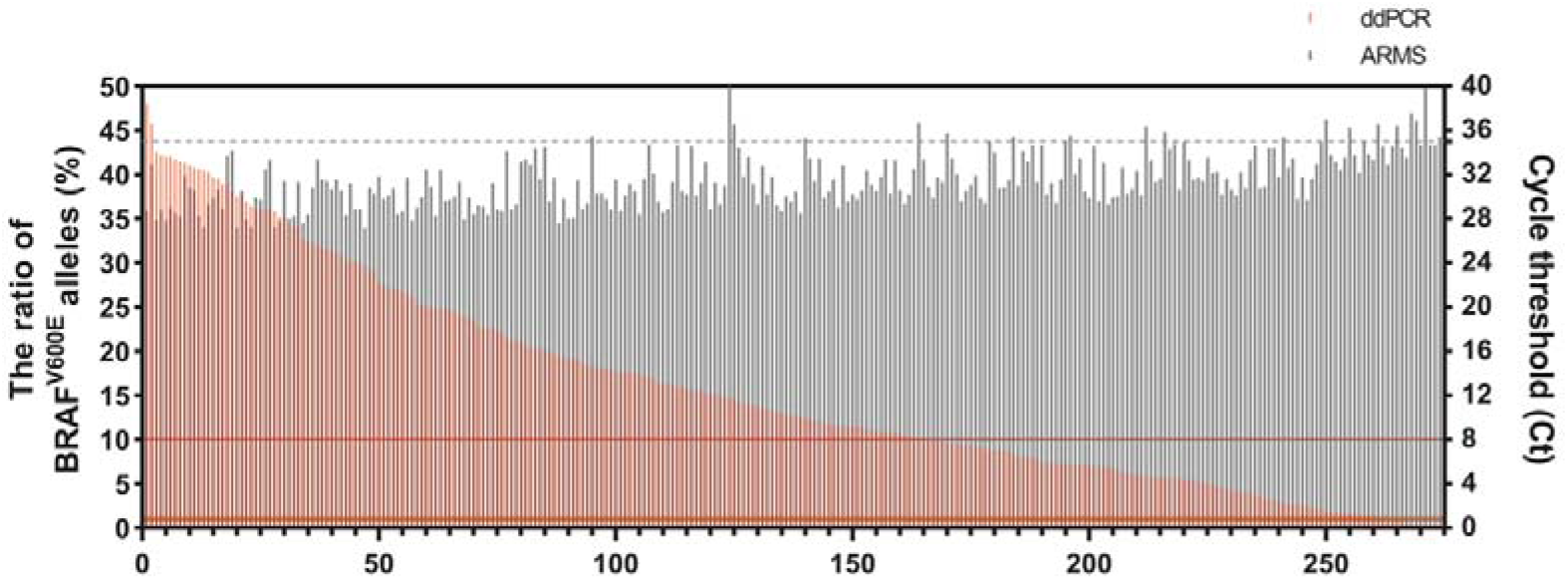
BRAFV600E mutation analysis in tissues sampled by ddPCR and ARMS in 275 PTC patients. The red bars indicate the ratio of BRAFV600E alleles detected by ddPCR (left Y-axis), in which 0.1% used as a cut-off value to define the BRAF-positive. The gray bars indicate the cycle threshold (Ct) detected by ARMS (right Y-axis). The red-thick line and fine line represent BRAFV600E alleles at ratio 1% and 10%, respectively. The gray dotted line indicates Ct value < 35 as a threshold to define the BRAF-positive by ARMS.

### Detection of the BRAFV600E mutation in PTC by ddPCR

#### Relationship between the ratio of BRAFV600E alleles and clinicopathological features of PTC

First, we analyzed the relationship between the BRAFV600E mutation status and clinicopathological features of PTC patients (Table 2). Second, we determined the relationship between the ratio of BRAFV600E alleles and clinicopathological features (Table 3). We found that the clinicopathological features of PTC patients were not closely related to the BRAFV600E mutation status (Table 2) but were closely correlated to the ratio of BRAFV600E alleles (Table 3). The ratio of BRAFV600E alleles was significantly associated with tumor size, capsule invasion/extrathyroidal extension, LNM, TNM stage, HT, and ATA recurrence risk. Sex, age and multifocality were independent of the ratio of BRAFV600E alleles.

**Table 2.**
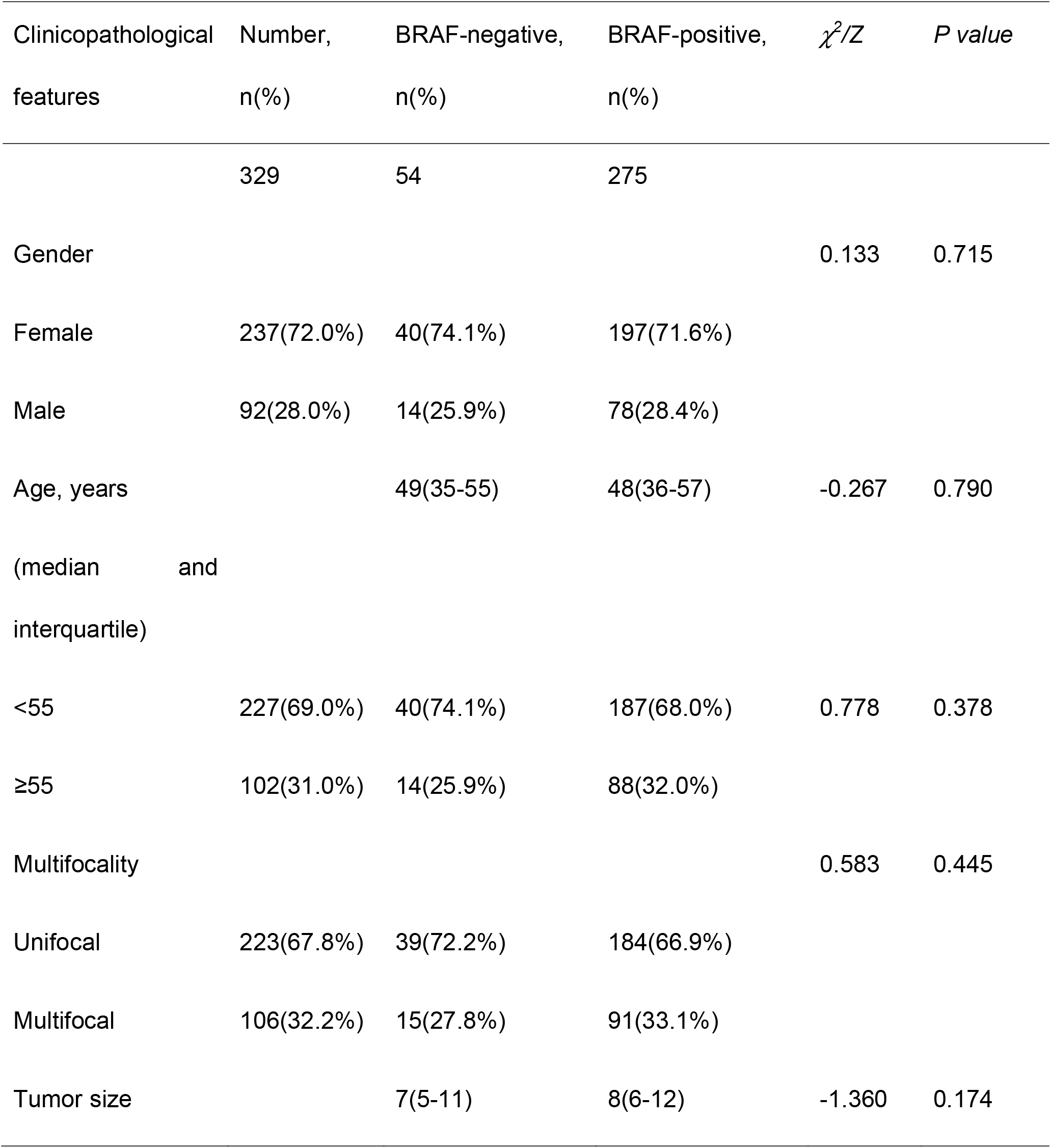
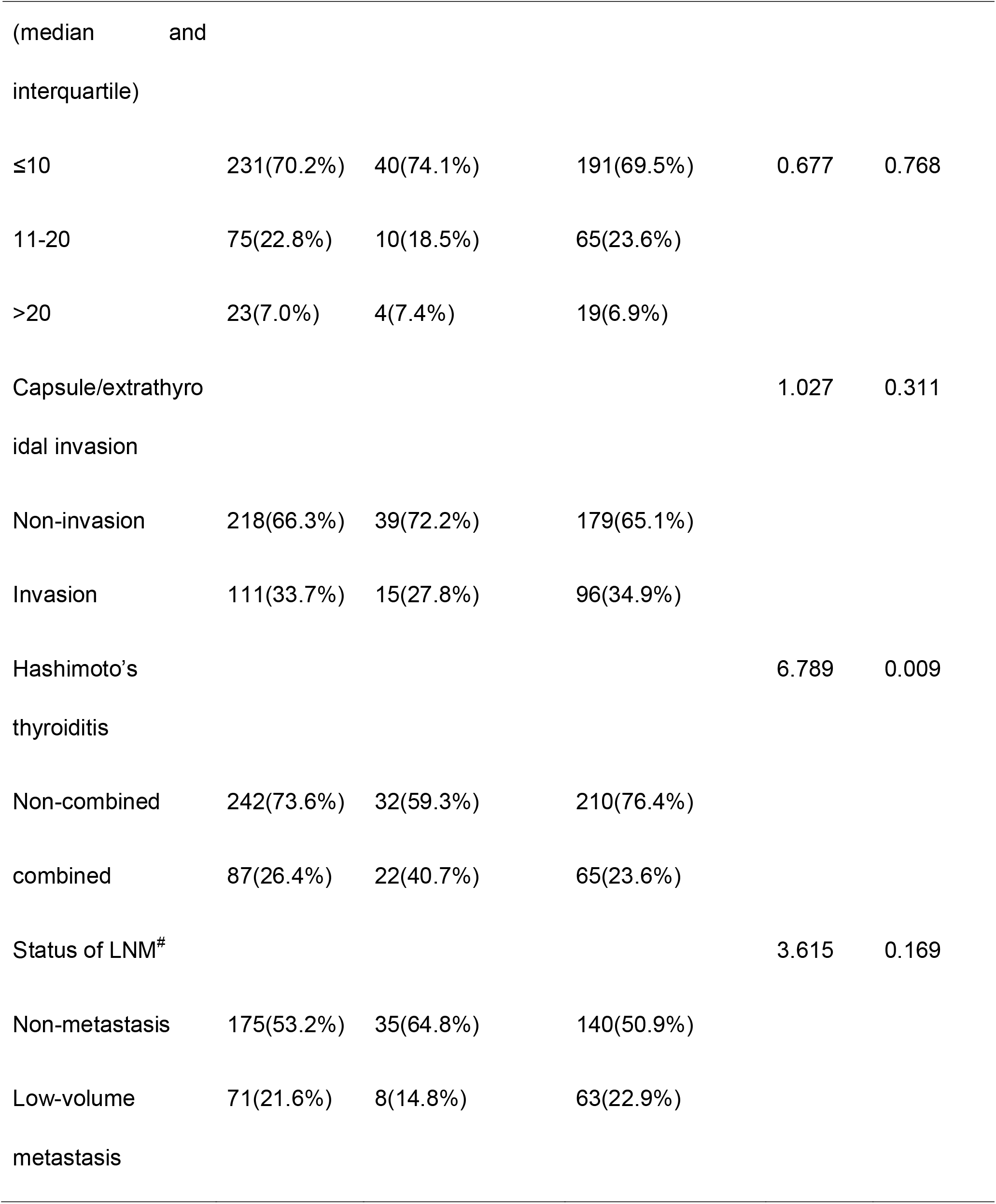
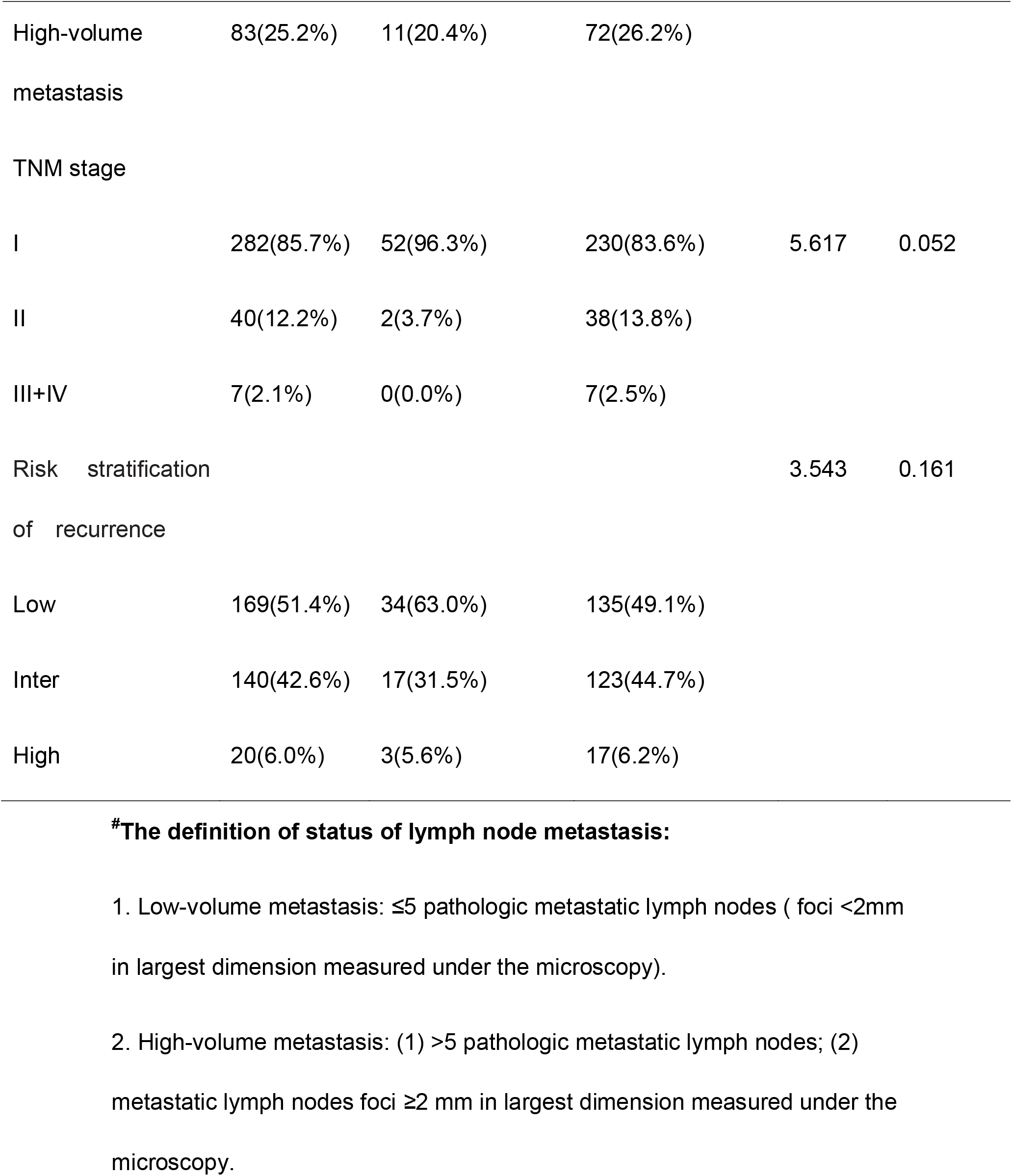
The relationship between BRAFV600E mutation status and clinicopathological features in PTC patients.

| Clinicopathological features | Number, n(%) | BRAF-negative, n(%) | BRAF-positive, n(%) | $\chi^2/Z$ | *P value* |
| --- | --- | --- | --- | --- | --- |
|  | 329 | 54 | 275 |  |  |
| Gender |  |  |  | 0.133 | 0.715 |
| Female | 237(72.0%) | 40(74.1%) | 197(71.6%) |  |  |
| Male | 92(28.0%) | 14(25.9%) | 78(28.4%) |  |  |
| Age, years<br>(median and interquartile) |  | 49(35-55) | 48(36-57) | -0.267 | 0.790 |
| <55 | 227(69.0%) | 40(74.1%) | 187(68.0%) | 0.778 | 0.378 |
| ≥55 | 102(31.0%) | 14(25.9%) | 88(32.0%) |  |  |
| Multifocality |  |  |  | 0.583 | 0.445 |
| Unifocal | 223(67.8%) | 39(72.2%) | 184(66.9%) |  |  |
| Multifocal | 106(32.2%) | 15(27.8%) | 91(33.1%) |  |  |
| Tumor size |  | 7(5-11) | 8(6-12) | -1.360 | 0.174 |
| (median | and |  |  |  |  |
| interquartile) |  |  |  |  |  |
| ≤10 | 231(70.2%) | 40(74.1%) | 191(69.5%) | 0.677 | 0.768 |
| 11-20 | 75(22.8%) | 10(18.5%) | 65(23.6%) |  |  |
| >20 | 23(7.0%) | 4(7.4%) | 19(6.9%) |  |  |
| Capsule/extrathyro |  |  |  | 1.027 | 0.311 |
| idal invasion |  |  |  |  |  |
| Non-invasion | 218(66.3%) | 39(72.2%) | 179(65.1%) |  |  |
| Invasion | 111(33.7%) | 15(27.8%) | 96(34.9%) |  |  |
| Hashimoto's |  |  |  | 6.789 | 0.009 |
| thyroiditis |  |  |  |  |  |
| Non-combined | 242(73.6%) | 32(59.3%) | 210(76.4%) |  |  |
| combined | 87(26.4%) | 22(40.7%) | 65(23.6%) |  |  |
| Status of LNM<sup>#</sup> |  |  |  | 3.615 | 0.169 |
| Non-metastasis | 175(53.2%) | 35(64.8%) | 140(50.9%) |  |  |
| Low-volume | 71(21.6%) | 8(14.8%) | 63(22.9%) |  |  |
| metastasis |  |  |  |  |  |
| High-volume | 83(25.2%) | 11(20.4%) | 72(26.2%) |  |  |
| metastasis |  |  |  |  |  |
| TNM stage |  |  |  |  |  |
| I | 282(85.7%) | 52(96.3%) | 230(83.6%) | 5.617 | 0.052 |
| II | 40(12.2%) | 2(3.7%) | 38(13.8%) |  |  |
| III+IV | 7(2.1%) | 0(0.0%) | 7(2.5%) |  |  |
| Risk stratification |  |  |  | 3.543 | 0.161 |
| of recurrence |  |  |  |  |  |
| Low | 169(51.4%) | 34(63.0%) | 135(49.1%) |  |  |
| Inter | 140(42.6%) | 17(31.5%) | 123(44.7%) |  |  |
| High | 20(6.0%) | 3(5.6%) | 17(6.2%) |  |  |

**#The definition of status of lymph node metastasis:**
1. Low-volume metastasis: $\leq 5$ pathologic metastatic lymph nodes ( foci $< 2\text{mm}$ in largest dimension measured under the microscopy). 2. High-volume metastasis: (1) $> 5$ pathologic metastatic lymph nodes; (2) metastatic lymph nodes foci $\geq 2\text{ mm}$ in largest dimension measured under the microscopy.

**Table 3.**
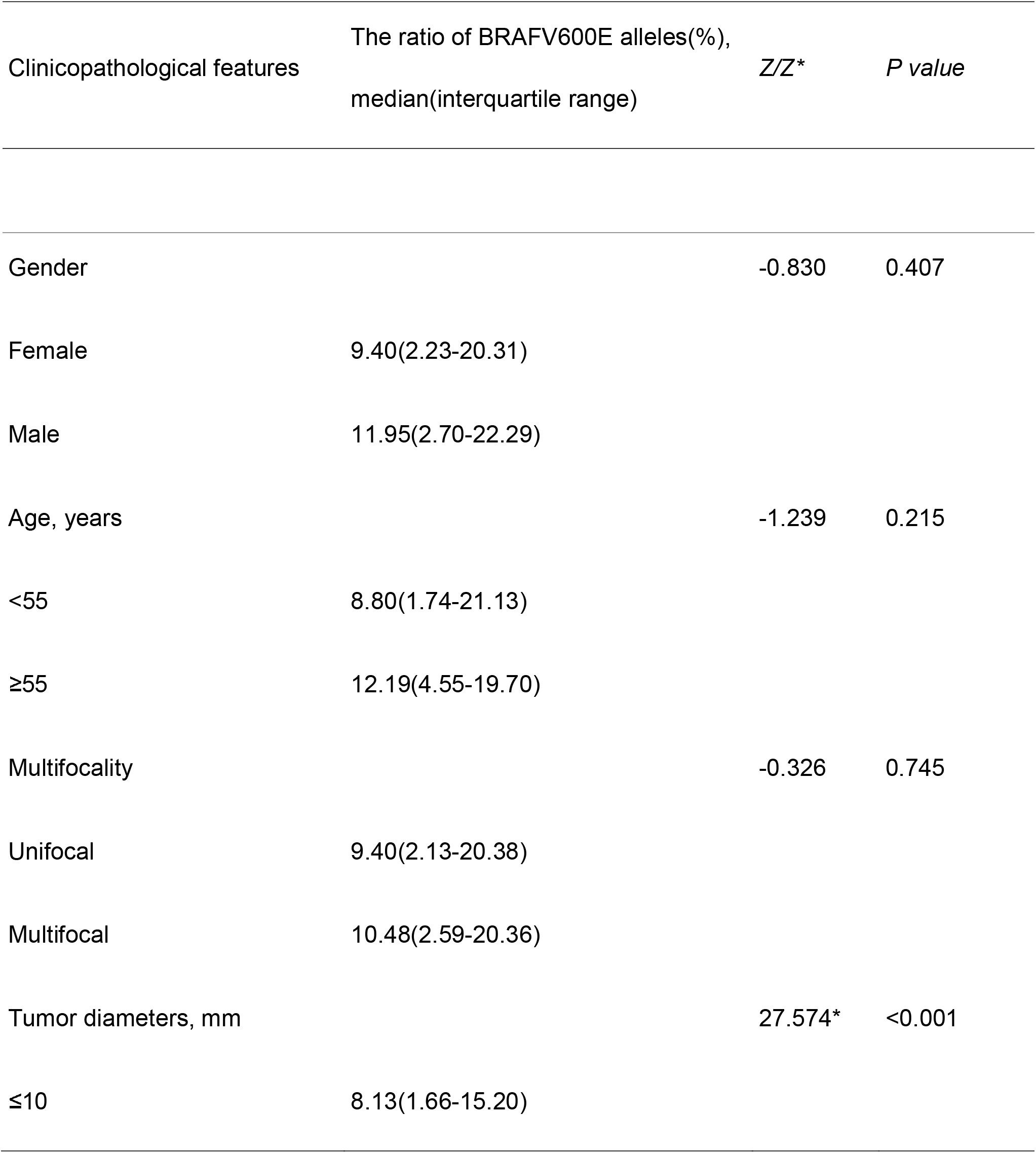
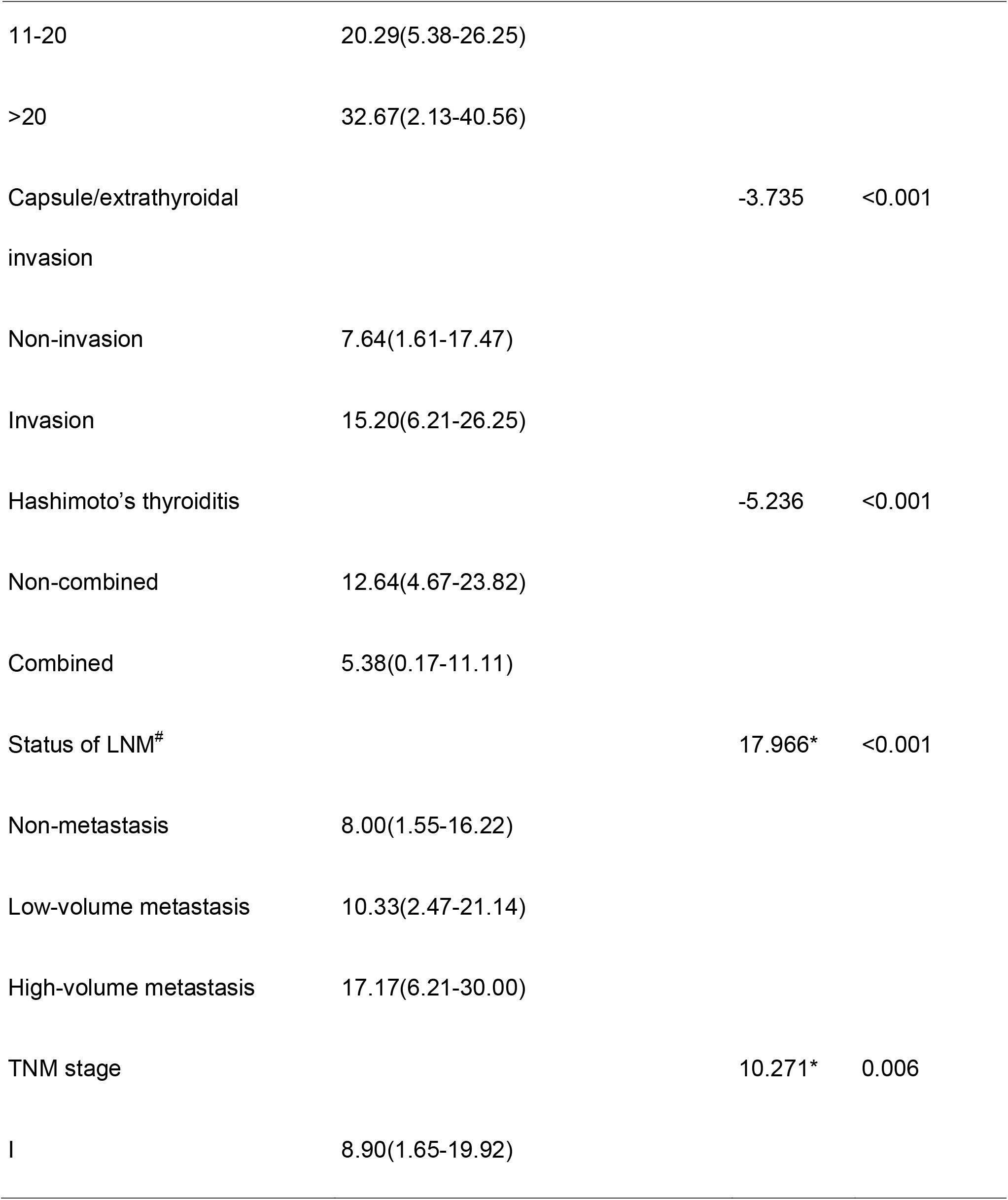
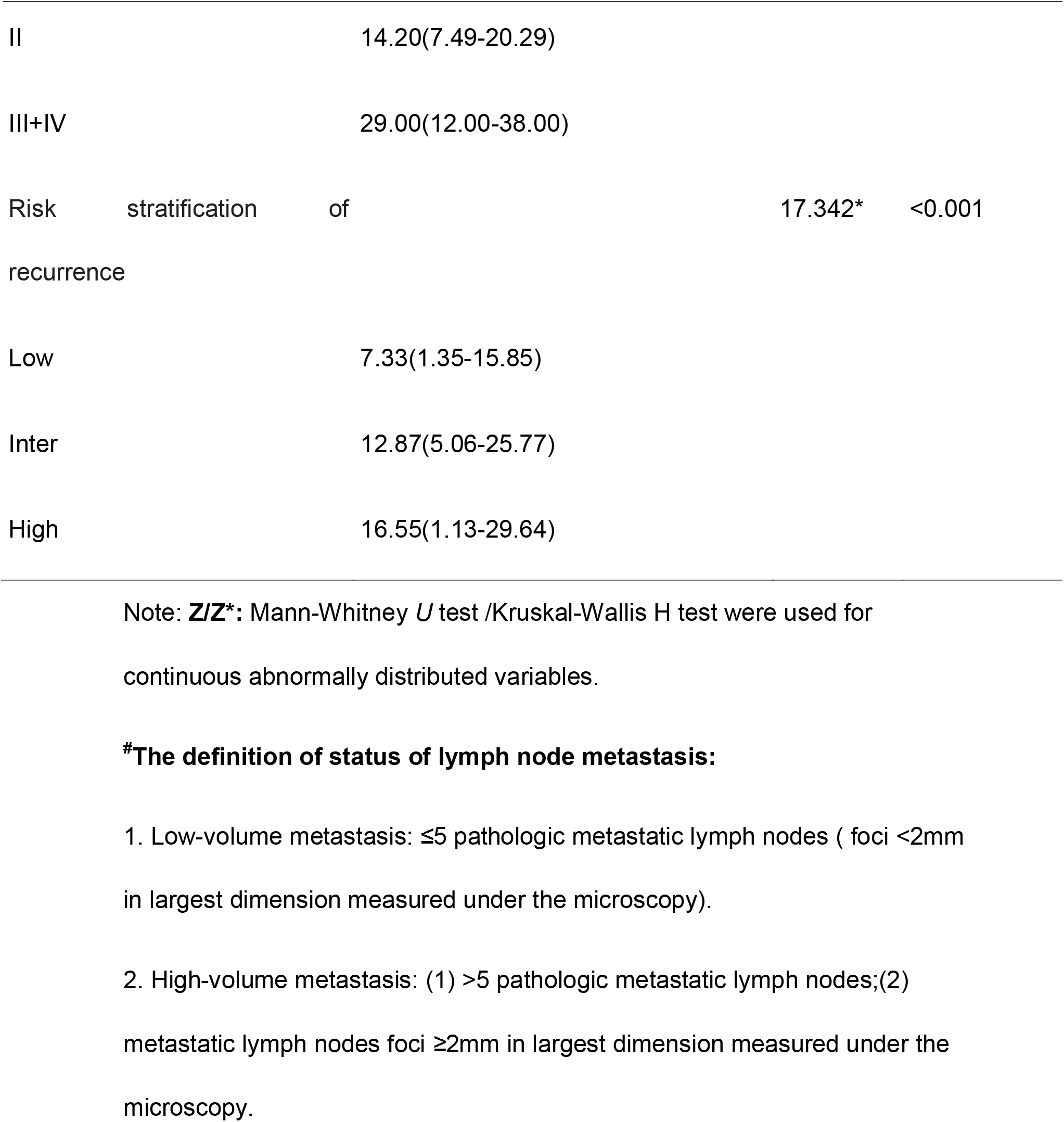
The relationship between the ratio of BRAFV600E alleles and clinicopathological features in PTC patients.

| Clinicopathological features | The ratio of BRAFV600E alleles(%),<br>median(interquartile range) | *Z/Z\** | *P value* |
| --- | --- | --- | --- |
| Gender |  | -0.830 | 0.407 |
| Female | 9.40(2.23-20.31) |  |  |
| Male | 11.95(2.70-22.29) |  |  |
| Age, years |  | -1.239 | 0.215 |
| <55 | 8.80(1.74-21.13) |  |  |
| ≥55 | 12.19(4.55-19.70) |  |  |
| Multifocality |  | -0.326 | 0.745 |
| Unifocal | 9.40(2.13-20.38) |  |  |
| Multifocal | 10.48(2.59-20.36) |  |  |
| Tumor diameters, mm |  | 27.574* | <0.001 |
| ≤10 | 8.13(1.66-15.20) |  |  |
| 11-20 | 20.29(5.38-26.25) |  |  |
| >20 | 32.67(2.13-40.56) |  |  |
| Capsule/extrathyroidal<br>invasion |  | -3.735 | <0.001 |
| Non-invasion | 7.64(1.61-17.47) |  |  |
| Invasion | 15.20(6.21-26.25) |  |  |
| Hashimoto's thyroiditis |  | -5.236 | <0.001 |
| Non-combined | 12.64(4.67-23.82) |  |  |
| Combined | 5.38(0.17-11.11) |  |  |
| Status of LNM<sup>#</sup> |  | 17.966* | <0.001 |
| Non-metastasis | 8.00(1.55-16.22) |  |  |
| Low-volume metastasis | 10.33(2.47-21.14) |  |  |
| High-volume metastasis | 17.17(6.21-30.00) |  |  |
| TNM stage |  | 10.271* | 0.006 |
| I | 8.90(1.65-19.92) |  |  |
| II | 14.20(7.49-20.29) |  |  |
| III+IV | 29.00(12.00-38.00) |  |  |
| Risk stratification of recurrence |  | 17.342* | <0.001 |
| Low | 7.33(1.35-15.85) |  |  |
| Inter | 12.87(5.06-25.77) |  |  |
| High | 16.55(1.13-29.64) |  |  |

Note: **Z/Z\***: Mann-Whitney *U* test /Kruskal-Wallis H test were used for continuous abnormally distributed variables.
**#The definition of status of lymph node metastasis:**
1. Low-volume metastasis: $\leq 5$ pathologic metastatic lymph nodes ( foci $< 2\text{mm}$ in largest dimension measured under the microscopy). 2. High-volume metastasis: (1) $> 5$ pathologic metastatic lymph nodes;(2) metastatic lymph nodes foci $\geq 2\text{mm}$ in largest dimension measured under the microscopy.

#### Relationship between the ratio of BRAFV600E alleles and maximal tumor diameter

In our cohort, the maximal tumor diameter ranged from 2 mm to 55 mm, with a median diameter of 8 mm. Patient samples were classified into three groups according to the maximal tumor diameter, ≤10 mm, 11-20 mm, and >20 mm, and their median ratios of BRAFV600E alleles were 8.13%, 20.29% and 32.67%, respectively. The ratio of BRAFV600E alleles was significantly different between these three groups (p<0.001) (Table 3). It appeared that there was a positive correlation between the ratio of BRAFV600E alleles and maximal tumor diameter (Figure 2). Thus, the ratio of BRAFV600E alleles increased with increasing maximal tumor diameter (Spearman’s rho=0.413, p<0.0001; R^2^=0.189, p<0.0001).

**Figure 2.**
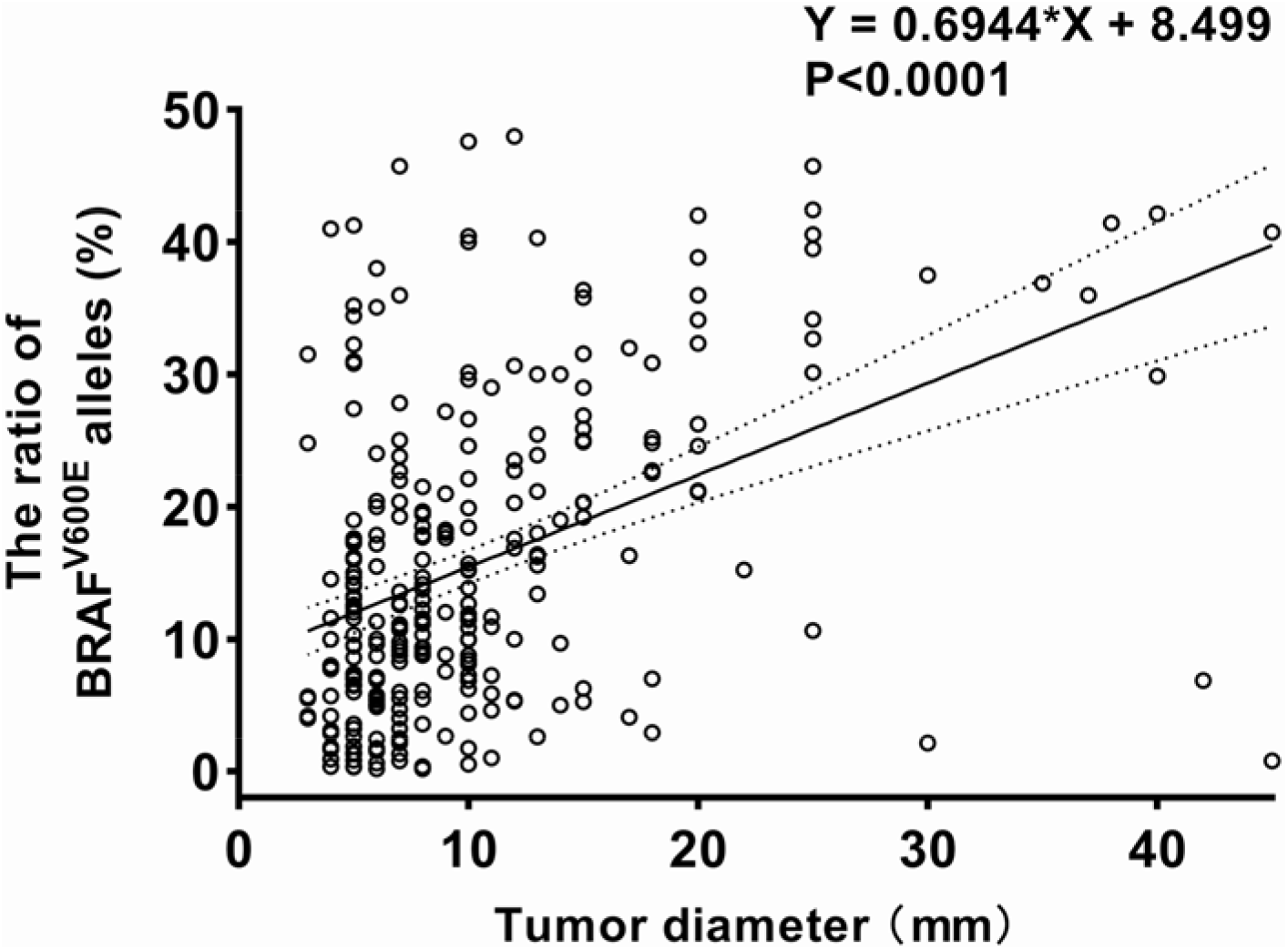
Correlation between BRAFV600E alleles ratio and maximal tumor diameter in PTC. Note: Scatter plot shows the correlation of the ratio of BRAFV600E alleles and tumor diameter. Lines represent simple linear regression ±95 % confidence interval. Spearman’s rho = 0.413, p<0.0001; simple regression adjusted R^2^ = 0.189, p<0.0001.

#### Relationship between the BRAFV600E allele ratio and capsule/extrathyroidal invasion

The median ratios of BRAFV600E alleles in 329 patients with and without thyroid capsule/extrathyroidal invasion were 15.20% and 7.64%, respectively. The ratios of BRAFV600E alleles were significantly different between these two groups of patients (p<0.001, Figure 3).

**Figure 3.**
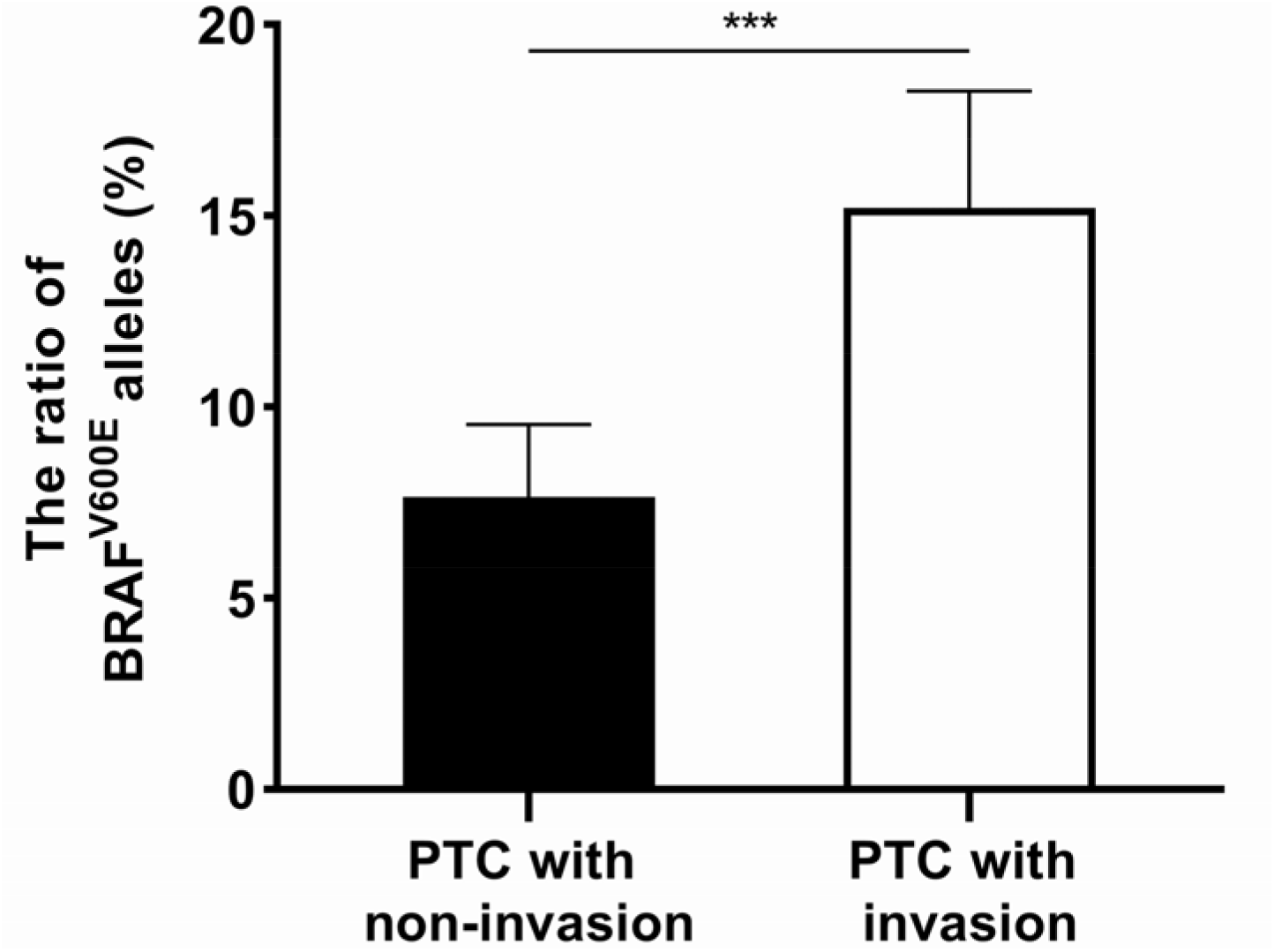
Comparison of BRAFV600E alleles ratio between capsule/extrathyroidal invasion and those without in PTC. Note: ***: P value <0.001; The top of columns represents the median, and lines represent ±95 % confidence interval.

#### Relationship between the BRAFV600E allele ratio and lymph node metastasis

A total of 154 patients had LNM, and 175 did not involve metastasis in our study. Among the patients with LNM, 71 were defined as having low-volume lymph node metastasis, with ≤5 pathologic metastatic lymph nodes and foci <2 mm in the largest dimension, and 81 were defined as having high-volume lymph node metastasis, with >5 pathologic metastatic lymph nodes or foci ≥2 mm in the largest dimension. The median ratios of BRAFV600E alleles were 8.00%,10.33% and 17.17%, respectively, in patients with no, low-volume and high-volume lymph node metastasis. The difference between these three groups was significant (p<0.001, Table 3). The ratio of BRAFV600E alleles was positively correlated with the number of metastatic lymph nodes (Spearman’s rho=0.220, p<0.0001; R^2^=0.070, p<0.0001, Figure 4) and the rate of lymph node metastasis (metastatic/dissected lymph nodes) (Spearman’s rho=0.232, p<0.0001; R^2^=0.058, p<0.0001, Figures 5).

**Figure 4.**
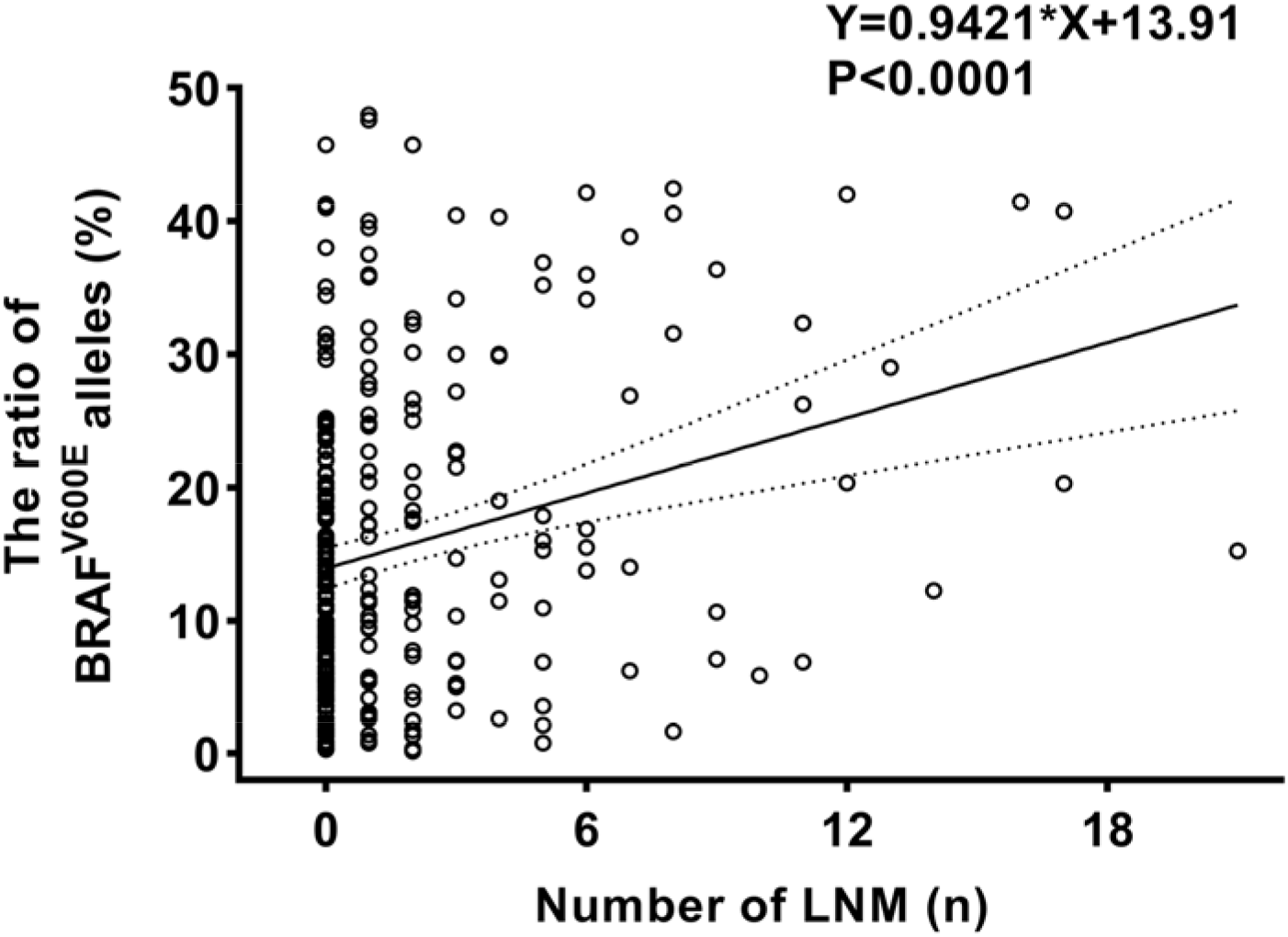
Correlation between BRAFV600E alleles ratio and the number of lymph node metastases in PTC. Note: Scatter plot shows the correlation of the ratio of BRAFV600E alleles and the number of LNM. Lines represent simple linear regression ±95% confidence interval. Spearman’s rho = 0.220, p<0.0001; simple regression adjusted R^2^ = 0.070, p<0.0001.

**Figure 5.**
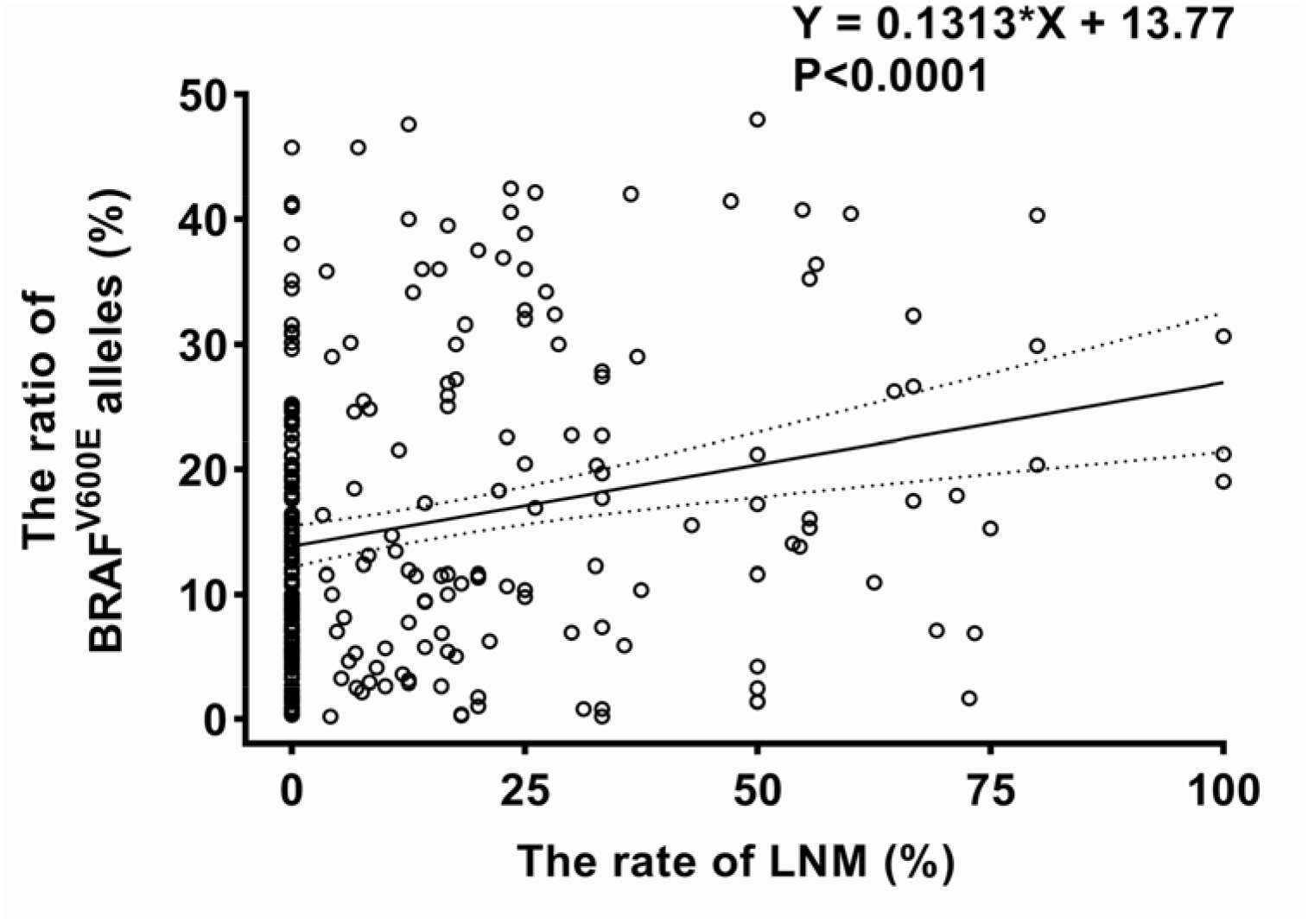
Correlation between BRAFV600E alleles ratio and the rate of lymph node metastases in PTC. Note: Scatter plot shows the correlation of the ratio of BRAFV600E alleles and the rate of LNM. Lines represent simple linear regression ±95% confidence interval. Spearman’s rho = 0.232, p<0.0001; simple regression adjusted R^2^ = 0.058, p<0.0001.

#### Relationship between the BRAFV600E allele ratio and Hashimoto’s thyroiditis

Eighty-seven PTC cases had accompanying HT. The median ratios of BRAFV600E alleles in PTC patients with and without HT were 5.38% and 12.64%, respectively, with a significant difference(p<0.001, Table 3, Figure 6). HT was most frequently observed in patients with stage I PTC than those with other stages of PTC. It was also noted that stage I PTC was associated with a low BRAFV600E mutation ratio (Table 4), suggesting that HT may has a negative impact on the BRAFV600E mutation in PTC.

**Table 4.** Comparison of BRAFV600E allele ratio and tumor stage between Hashimoto’s thyroiditis and those without in PTC patients.

| Hashimoto's<br>thyroiditis | TNM stage, n(%) | | $\chi^2$ | *P value* | The ratio of BRAFV600E<br>alleles | *P value* |
| --- | --- | --- | --- | --- | --- | --- |
|  | Stage I | Stage II-III-IV |  |  |  |  |
| Non-combined | 201(83.1%) | 41(16.9%) | 5.274 | 0.022 | 12.64(4.67-23.82) | <0.001 |
| Combined | 81(93.1%) | 6(6.9%) |  |  | 5.38(0.17-11.11) |  |

**Figure 6.**
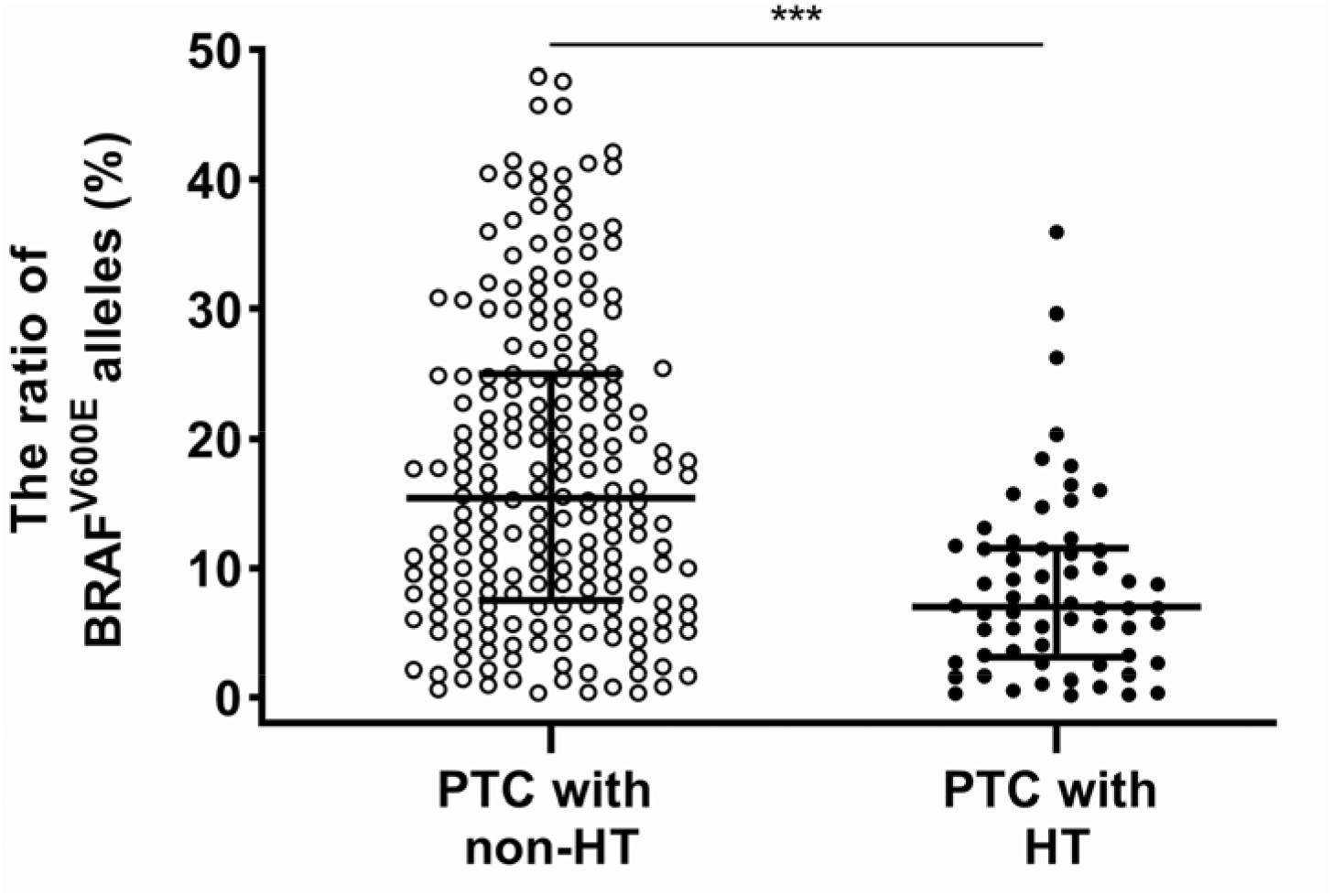
Comparison of BRAFV600E alleles ratio between Hashimoto’s thyroiditis and those without in PTC. Note: ***: P value <0.001. Vertical scatter plots of the ratio of BRAFV600E alleles grouped by PTC combined Hashimoto’s thyroiditis. Lines represent median and interquartile range.

#### Relationship between the BRAFV600E allele ratio and tumor stage

According to the 8th revision of the TNM staging system by the AJCC, there were 282 patients with stage I disease, 40 patients with stage II disease, and 7 patients with stage III or IV disease among all 329 patients, and their median ratios of the BRAFV600E alleles were 8.90%, 14.20% and 29.00%, respectively, with significant differences(p=0.006). We found that the BRAFV600E mutation ratio increased as the tumor advanced from stage I to stage IV (Table 3).

#### Relationship between the BRAFV600E allele ratio and tumor recurrence risk

We analyzed the postoperative clinicopathological characteristics of all PTC patients according to the 2015 ATA guidelines for the risk stratification standard of thyroid cancer recurrence. As a result, 169 patients were considered low risk, 140 were considered intermediate risk, and 20 were considered high risk. The median ratios of the BRAFV600E alleles were 7.33%, 12.87% and 16.55% for low-, intermediate- and high-risk patients, respectively, with a significant difference (p<0.001). Moreover, the ratio of the BRAFV600E alleles increased as the risk of tumor recurrence increased (Table 3).

## DISCUSSION

Although the BRAFV600E mutation has been linked to the occurrence and development of PTC and associated with its clinicopathological characteristics since 2003, the results are still debatable. Several factors contribute to the inconsistent data. First, the different detection methods used can result in wide variations in the rate of BRAF mutations found in PTC. For example, the positive rate of BRAFV600E detected by NGS in PTC is usually over 80%(33), whereas that detected by Sanger sequencing is approximately 45-59%(34). Currently, most studies use a qualitative analysis rather than a quantitative analysis of BRAFV600E mutations. Second, the difference in the number of cases analyzed may lead to various outcomes. Gandolfi et al studied 132 cases of PTC and concluded that the occurrence and percentage of the BRAFV600E mutated allele did not have an obvious role in the development of PTC metastases and that the average mutated allele percentage decreased as PTC progressed from the primary location to sites of lymph node metastasis(35). Thus, their results suggest that the less frequent the BRAFV600E mutation is, the more aggressive the tumor, which opposes the common concept of the pathologic function of BRAFV600E in PTC(36,37). It is generally believed that results from a small number of cases are not reliable. Third, the difference may be caused by the analysis of different clinicopathological indexes. Previous studies included the tall cell and follicular subtypes with traditional PTC and found that BRAFV600E mutations were exactly related to tumor aggressiveness. However, if nontraditional PTC was excluded, no correlation was found(34,38,39). Therefore, unified, traditional PTC must be included to assess its correlation with BRAFV600E mutations. Finally, different disease stages may also contribute to the variation. For example, Gan et al. reported that BRAFV600E was closely associated with the clinicopathological characteristics of patients with stage II or above PTC, but such an association was not observed in patients with stage I PTC(40). Therefore, the difference in the proportion of PTC patients in different tumor stages may also contribute to the variation. In this study, we employed the most updated detection technology to quantitatively analyze BRAFV600E mutations in 329 patients with traditional PTC. To the best of our knowledge, this is the largest number of PTC cases studied so far. We selected a total of 11 clinicopathological characteristics for the analysis, the most comprehensive and greatest number of indexes analyzed thus far.

This study found that the sensitivity of ddPCR to detect BRAFV600E mutations in PTC was almost equal to that of NGS, and higher than that of ARMS (Table1). The BRAFV600E mutation was undetectable by ARMS but detectable by ddPCR in 13 patients. Among the 329 PTC patients, the ratio of BRAFV600E alleles in 165 cases was less than 10%, which is usually beyond the range detected by the first-generation gene detection technology(26,41).

The aggressiveness of PTC is usually determined by tumor size, capsule and extrathyroidal invasion, LNM, and pulmonary and bone metastasis. These clinicopathological features of PTC can directly indicate the tumor stage and objectively reflect the risk of tumor recurrence. These indexes have been consistently recognized and used to guide the clinical diagnosis and treatment of PTC. In our study, the BRAFV600E allele ratio but not the BRAFV600E mutation in PTC tissues was positively correlated with these features. Therefore, the aggressiveness of PTC can be reliably assessed by the quantitative index of the BRAF600E allele ratio, making the BRAFV600E allele ratio a molecular marker for the clinical evaluation of PTC aggressiveness.

The detection of the BRAFV600E allele ratio can be influenced by the coexistence of other pathologic conditions in the thyroid. The study by Colombo et al indicated that some wild-type BRAF alleles detected in PTC were in fact from inflammatory or stromal cells, regarded as “contamination”(42). Accordingly, the frequency of mutated alleles must be normalized to the percentage of neoplastic cells in the samples(43). After the exclusion of contamination, we found significant differences in the BRAFV600E mutation ratio in the whole group of PTC, which presented different biological behaviors and clinical characteristics.

It is well known that a proportion of PTC can coexist with or develop from HT, an autoimmune disease. In this study, we stratified PTC patients with or without HT and found that PTC patients with HT had a lower BRAFV600E allele ratio than those without. Moreover, the proportion of stage I PTC patients with HT was significantly higher than that of those without (Table 4). These observations are supported by the fact that the lymphocytic infiltration of HT is a protective factor against PTC progression(44). This clinical phenomenon can also be explained by another finding in our study: PTC patients with HT carried fewer BRAFV600E mutations are likely to be less aggressive. However, the mechanism responsible is unclear. It is possible that inflammation in the thyroid may prevent the occurrence of BRAFV600E mutations or have a negative impact on BRAFV600E mutations.

The risk stratification of PTC recurrence recommended by the ATA in 2015 has important clinical guiding value for postoperative iodine-131 treatment and follow-up(27). In our study, the median ratio of BRAFV600E alleles in low-, intermediate- and high-risk patients was 7.33%, 12.87%, 16.55%, respectively, with a significant difference (p<0.001). It was noted that the size of the tumor may not always be positively correlated with the ratio of BRAFV600E alleles. For example, in the high-risk group, the tumor diameter of some patients was only 6 mm, but the BRAFV600E allele ratio was up to 14.3%. However, in the low-risk group, the tumor diameter of some patients reached 12 mm, while the BRAFV600E allele ratio was only 0.2%. Therefore, it is more accurate or meaningful to evaluate the risk of recurrence of PTC by quantitatively detecting the BRAF allele ratio rather than measuring the tumor diameter. Furthermore, if quantitative detection of the BRAF allele ratio is applied in a sample obtained from preoperative FNA, this concept and technique will provide important guidance for decisions regarding the management of PTC, including the pattern of thyroidectomy and cervical lymph node dissection.

Several limitations to this study should be acknowledged. First, in this study, 83.58% of PTC patients had the BRAFV600E mutation, and 16.42% did not, indicating that the BRAFV600E mutation is not the only factor contributing to the occurrence and development of PTC. Though the BRAFV600E mutation is the most frequent genetic event found in PTC, other genetic changes such as TERT, KRAS and HRAS mutations, RET fusions, and TRK rearrangements have also been reported(42,45). Therefore, an accurate risk stratification of PTC requires not only a mutation analysis of a single genetic event, but also a comprehensive analysis of other types of genetic alterations. Second, parts of the tumor tissues subjected to genetic testing and staining may not be matched exactly. Third, since intratumor heterogeneity is objective, tumor tissues subjected to mutation detection may not be representative of the whole tumor(46). To the best of our knowledge, the sample number we examined was the largest so far, but selection bias, which affects the detection results, cannot be excluded. Finally, our work is a single-center retrospective study, and a multicenter prospective study is required to further evaluate the prognostic significance of the ratio of BRAFV600E alleles in PTC.

## CONCLUSIONS

ddPCR can accurately and quantitatively detect the BRAFV600E allele ratio in PTC. The BRAFV600E allele ratio is closely related to the clinical aggressiveness of PTC and can reflect the biological behavior of PTC. Therefore, it can be used as a molecular-based stratification index of recurrence risk and should aid in decisions regarding the clinical diagnosis and treatment of PTC.

## Data Availability

Data that support the findings of this report are available from the corresponding author upon request.

## ACKNOWLEDGMENTS

This work was supported by grant from the Key Project of Scientific and Technological Innovation in Hangzhou (award number: 20131813A08) and the Medical Science Research Program of Hangzhou (award number: OO20190490). The authors are grateful to all members of the Department of Surgical Oncology, Centre of Translational Medicine, and Department of Pathology at Hangzhou First People’s Hospital.

## AUTHORS CONTRIBUTIONS

DC.L and YQ.N designed the study and wrote the paper. YP.X, P.Z, JJ.S and S.L analyzed the data and performed the study. F.W, TH.Z, SH.S and KN.L contributed with data collection and patients follow-up. DC.L and SR.Z supervised the whole process and critically revised the paper. All authors read and approved the final version.

