## Supplemental table 1 for "The ratio of BRAFV600E alleles can be used to assess the biological behavior of papillary thyroid carcinoma"

**Supplemental table 1. The result of BRAFV600E mutation in 16 PTCs detected by ddPCR, ARMS and NGS**

| No. | ddPCR result | ARMS result | NGS result | Final result^#^ |
| --- | --- | --- | --- | --- |
| 1 | P | S | P | P |
| 2 | P | S | p | p |
| 3 | P | S | P | P |
| 4 | P | S | P | P |
| 5 | P | S | P | P |
| 6 | P | S | P | P |
| 7 | N | S | N | N |
| 8 | P | S | P | P |
| 9 | P | S | P | P |
| 10 | P | S | P | P |
| 11 | P | N | P | P |
| 12 | P | N | P | P |
| 13 | N | P | N | N |
| 14 | N | P | P | P |
| 15 | P | S | P | P |
| 16 | P | S | P | P |

Abbreviation: P=Positive; N=Negative; S=Suspicious

^#^ The definition of the final result: among the three detection result, two of the consistent results was considered the final results.
